# Association of sleep abnormalities in older adults with risk of developing Parkinson’s disease

**DOI:** 10.1101/2022.05.08.22274816

**Authors:** Abidemi I. Otaiku

**Affiliations:** Department of Neurology, Birmingham City Hospital, Birmingham, UK; Centre for Human Brain Health, University of Birmingham, Birmingham, UK

## Abstract

**Background:** Parkinson’s disease (PD) is associated with abnormalities of sleep macro- and microstructure as measured using polysomnography (PSG). Whether sleep abnormalities precede the development of PD is unknown. This study investigated the association between PSG measured sleep abnormalities in older adults and the risk of developing PD.

**Methods:** 2,770 men aged 67 years or older from the ancillary sleep study of the Osteoporotic Fractures in Men Study (MrOS), a population-based cohort from the USA, who were free from PD at baseline (December 2003 – March 2005) and in whom there was PSG data available, were included in this analysis. Incident PD was based on a clinical diagnosis from a medical professional. Multivariable logistic regression was used to estimate odds ratios (OR) for incident PD by quartiles of PSG measures.

**Findings:** During a median follow-up of 10.1 years, 70 (2.5%) cases of incident PD were identified. Longer total sleep time, lower rapid eye movement sleep (REM) percentage, a lower α/θ ratio during non-REM sleep and higher minimum oxygen saturations during REM sleep, were independently associated with an increased risk of developing PD during follow-up. Conversely, a higher awakening index was independently associated with a decreased risk of developing PD. The OR for the highest risk quartiles compared to the lowest risk quartiles, ranged from 2.3 to 4.0 (*P* < 0.05). The associations remained significant when incident PD cases occurring within the first two years of follow-up were excluded from the analyses.

**Conclusions:** Macro- and microstructural sleep abnormalities precede the development of PD by several years and can identify individuals at high risk of developing PD in the future. Future studies are needed to determine whether sleep abnormalities represent preclinical markers of PD or causal risk factors.

## Introduction

Parkinson’s disease (PD) is the second most common neurodegenerative disease and the fastest growing neurological disorder in the world.^1^ Alongside the characteristic motor features of PD, a significant proportion of people with PD also experience sleep-wake disturbances. In particular, 40-90% of people with PD have at least one sleep disorder,^2^ and certain sleep disorders - such as rapid eye movement sleep behaviour disorder (RBD) - can predate the onset of PD by several years or even decades.^3^

Although it is well-known that people with PD exhibit alterations in their sleep macrostructure (routine polysomnographic [PSG] characteristics)^4^ and microstructure (quantitative analysis of the sleep electroencephalogram [qEEG]),^5^ it remains unknown whether these alterations might precede the development of PD.

Establishing the temporal relationship between objective sleep alterations and the development of PD, may have important value for early diagnosis, better understanding the pathophysiology of PD, and might open up new approaches for delaying or preventing PD onset.

This study investigated the hypothesis that macro- and microstructural sleep alterations precede the development of PD, and sought to test this theory using data from the Outcomes of Sleep Disorders in Older Men (MrOS) Sleep Study.^6^

## Methods

### Study design and participants

MrOS is an observational, longitudinal cohort study that enrolled 5994 community-dwelling men aged 65 years and over at six clinical centres in the United States, including: Birmingham, Minneapolis, Palo Alto, Pittsburgh, Portland and San Diego.^6^ The MrOS Sleep Study is an ancillary project that included 3135 men who were recruited from December 2003 to March 2005, and underwent comprehensive sleep assessments. Details regarding the studies have been described in detail elsewhere.^6,7^

The sample for the current analysis was limited to participants who were free from PD at the baseline sleep visit, had no missing data for PSG, qEEG, self-reported sleep measures, or any covariates, and took part in at least one follow-up after baseline. This left a final analytic sample of 2,770 participants.

### Sleep macrostructure

A single-night, unattended, portable, in-home PSG (Safiro; Compumedics) was conducted at baseline as previously described.^8^ The recording montage included: C3-A2 and C4-A1 EEGs, bilateral electrooculogram (EOG), bipolar submental electromyogram (EMG); thoracic and abdominal respiratory inductance plethysmography, airflow (detected by a nasal-oral thermistor and nasal pressure cannula), finger pulse oximetry (SpO2), bilateral leg movements (using piezoelectric sensors), and electrocardiogram (ECG).

Routine PSG characteristics were evaluated based on previously published definitions.^7,8^ The measures used in the present study included: total sleep time (TST), percentage of TST spent in stage 1, stage 2, slow-wave sleep (SWS; stages 3 and 4 combined) and rapid eye movement (REM) sleep; awakening index (number of awakenings/hour), apnoea-hypopnea index (AHI; accompanied by a 3% or greater oxygen desaturation), minimum oxygen saturations (min SpO2), and periodic limb movement index (PLMI).

### Sleep microstructure (qEEG)

The EEG signals from the PSG recordings were subjected to an off-line spectral analysis using a Fast-Fourier Transform Routine.^9^ The absolute and relative power of the EEG in different frequency bands throughout the night were computed for REM and non-REM (NREM) separately. In this analysis, the qEEG measures of interest included the mean relative power for the following frequency bands: δ (0.1 – 4 Hz), θ (4 – 8 Hz), α (8 – 12 Hz), β (12 – 30 Hz) and γ (>30 Hz). In addition, using the absolute spectral power for α and θ, the α/θ power ratio (α/θ) - a synoptic index of EEG background slowing down - was calculated.^10^

### Self-reported sleep parameters

All participants completed the Pittsburgh Sleep Quality Index (PSQI) at baseline, a validated questionnaire for assessing habitual sleep quality and disturbances.^11^ Self-reported estimates for habitual TST and awakenings over the past month were included in this analysis as secondary exposures. Habitual TST was assessed using responses to PSQI item 4: “During the past month, how many hours of actual sleep did you get at night? (This may be different than the number of hours you spent in bed)”. Habitual awakening frequency was assessed using item 5b: “During the past month, how often have you had trouble sleeping because you wake up in the middle of the night or early morning?”.

The question for TST included a free text response, and the question for awakenings included the following options: (1) Not during the past month, (2) Less than once a week, (3) Once or twice a week, and (4) Three or more times a week.

### Ascertainment of incident PD

During the 12-year follow-up period, participants were asked at four clinical visits and one questionnaire-based contact, to report whether they had ever been diagnosed with PD by a physician. Incident PD was defined as physician-diagnosed PD. Follow-up time was calculated for each participant as the time to the most recent collection of data on PD diagnosis.

### Covariates

Covariates that might confound the association between PSG characteristics and incident PD in this analysis, included: age in years (continuous), ethnicity (white, non-white), educational qualifications (college degree/high school degree/none), smoking status (current, past, never), history of physician-diagnosed diabetes mellitus (yes/no), history of physician-diagnosed hypertension (yes/no), depressive symptoms (continuous), global cognitive function (continuous), daytime sleepiness (continuous), physical activity levels (continuous), body mass index (continuous), caffeine intake (continuous), and use of psychotropic medications (yes/no).

Age, ethnicity, educational qualifications, smoking status, history of physician diagnosed diabetes mellitus and history of physician diagnosed hypertension, were self-reported at baseline. Depressive symptoms were evaluated using the Geriatric Depression Scale (scores range from 0-15, with higher scores indicating more severe depressive symptoms).^12^ Global cognitive function was measured using The Modified Mini-Mental State Examination (scores range from 0-100, with higher scores indicating better cognitive function).^13^ Daytime sleepiness was assessed using The Epworth Sleepiness Scale (scores range from 0-24, with higher scores indicating increased daytime sleepiness).^14^ Level of physical activity was estimated using the Physical Activity Scale for the Elderly (scores range from 0-793, with higher scores indicating greater physical activity levels).^15^ Body mass index was calculated as weight divided by height (kg/m2). Caffeine intake (mg/day) was calculated using responses to survey questions on habitual intake of coffee, tea, and soda.^16^ Psychotropic medication use was defined as: antidepressants, benzodiazepines, antipsychotics, stimulants, anticonvulsants, dementia medications, nonbenzodiazepine, nonbarbiturate sedative hypnotics and melatonin.

### Statistical analysis

Characteristics of the participants at baseline were compared by incident PD status using χ^2^ tests for categorical variables, independent samples t-tests for normally distributed continuous variables, and Mann–Whitney U tests for nonnormally distributed variables.

The associations of macro- and microstructural sleep characteristics with incident PD were assessed using logistic regression to determine odds ratios (OR) with 95% confidence intervals (CI). All PSG and qEEG measures were expressed as quartiles. The risk of incident PD for each quartile was compared with the reference quartile, and a *P* value for the trend across quartiles was determined by entering quartile into the model as continuous variable. Model 1 was minimally adjusted for age and clinic site. Model 2 additionally adjusted for ethnicity, education, smoking status, diabetes, hypertension, depressive symptoms, cognitive function, daytime sleepiness, physical activity levels, body mass index, caffeine intake and psychotropic medication use,

Several sensitivity analyses were performed to confirm the robustness of the findings. The analyses for the macro- and microstructural sleep measures found to be significantly associated with PD in the fully adjusted models, were repeated after: (1) adjusting for all the other measures found to be associated with PD (to determine whether the measures were independent of one another), (2) introducing a lag time of approximately 2 years, including only PD cases identified after the first follow-up visit (to minimise the possibility of reverse causality), and (3) adjusting for the presence of physician-diagnosed non-apnoea sleep disorders (to assess whether the results were driven by participants with primary sleep pathologies, e.g. idiopathic RBD). A sensitivity analysis was carried out for the sleep-related breathing measures (AHI, min SpO2), which further adjusted for pre-sleep resting oxygen saturations (to assess whether the results were affected by underlying lung impairment). Finally, self-reported habitual TST and awakening frequency were assessed as secondary exposures (to determine whether the predictive value of self-reported sleep characteristics would be congruent with objectively measured characteristics).

Statistical testing was performed two-sided at *P* <0.05. All analyses were performed using SPSS version 28 (IBM Corp., Armonk, NY).

## Results

Demographics and baseline sleep measures of the participants stratified by incident PD status, are presented in Table 1 and Table 2 respectively. Among 2,770 participants included in this analysis, 70 cases of incident PD (2.5%) were identified during a median follow-up of 10.1 years. Of these cases, 10% occurred within the first 2 years of follow-up, and 90% occurred between 2 and 12.2 years.

**Table 1.** Sample characteristics at baseline by incident PD status.

| Characteristic | No PD<br>(n = 2700) | Incident PD<br>(n = 70) | *P*<br>value |
| --- | --- | --- | --- |
| Age (years) | 76.3 ± 5.5 | 76.1 ± 5.3 | 0.83 |
| White, n (%) | 2479 (91.8) | 68 (97.8) | 0.11 |
| Education, n (%) |  |  | 0.36 |
| College degree | 1522 (56.4) | 45 (64.3) |  |
| High school degree | 1036 (38.4) | 21 (30.0) |  |
| No qualifications | 142 (5.3) | 4 (5.7) |  |
| Smoking, n (%) |  |  | 0.83 |
| Current | 55 (2.0) | 2 (2.9) |  |
| Past | 1577 (58.5) | 39 (55.7) |  |
| Never | 1068 (39.6) | 29 (41.4) |  |
| Alcohol intake, drinks/wk, n (%)<sup>a</sup> |  |  | 0.39 |
| <1 | 1249 (46.4) | 29 (42.0) |  |
| 1-13 | 1290 (48.0) | 38 (55.1) |  |
| ≥14 | 150 (5.6) | 2 (2.9) |  |
| Diabetes, n (%) | 355 (13.1) | 6 (8.6) | 0.26 |
| Hypertension, n (%) | 1354 (50.1) | 36 (51.4) | 0.83 |
| Non-apnoea sleep disorder, n (%) |  |  | 0.96 |
| Yes | 135 (5.0) | 4 (5.7) |  |
| No | 2523 (93.4) | 65 (92.9) |  |
| Unknown | 42 (1.6) | 1 (1.4) |  |
| Depressive symptoms score<sup>b</sup> | 1.7 ± 2.1 | 2.6 ± 2.6 | <0.001 |
| Cognitive function score<sup>c</sup> | 92.8 ± 6.2 | 92.4 ± 4.6 | 0.09 |
| Daytime sleepiness score<sup>d</sup> | 6.1 ± 3.6 | 6.1 ± 4.2 | 0.65 |
| Physical activity score<sup>e</sup> | 146.6 ± 71.2 | 151.7 ± 73.1 | 0.54 |
| BMI (kg/m<sup>2</sup>) | 27.2 ± 3.8 | 27.5 ± 3.4 | 0.15 |
| Caffeine intake (mg/day) | 237.9 ± 245 | 251.0 ± 273 | 0.83 |
| Psychotropic medication use, n (%) | 438 (16.2) | 19 (27.1) | 0.02 |

Plus-minus values are means ± SD
a $n = 2,689/69$
b Scores range from 0-15, with higher scores indicating more severe depressive symptoms
c Scores range from 0-100, with higher scores indicating better cognitive function
d Scores range from 0-24, with higher scores indicating increased daytime sleepiness
e Scores range from 0-793, with higher scores indicating greater physical activity levels

**Table 2.**
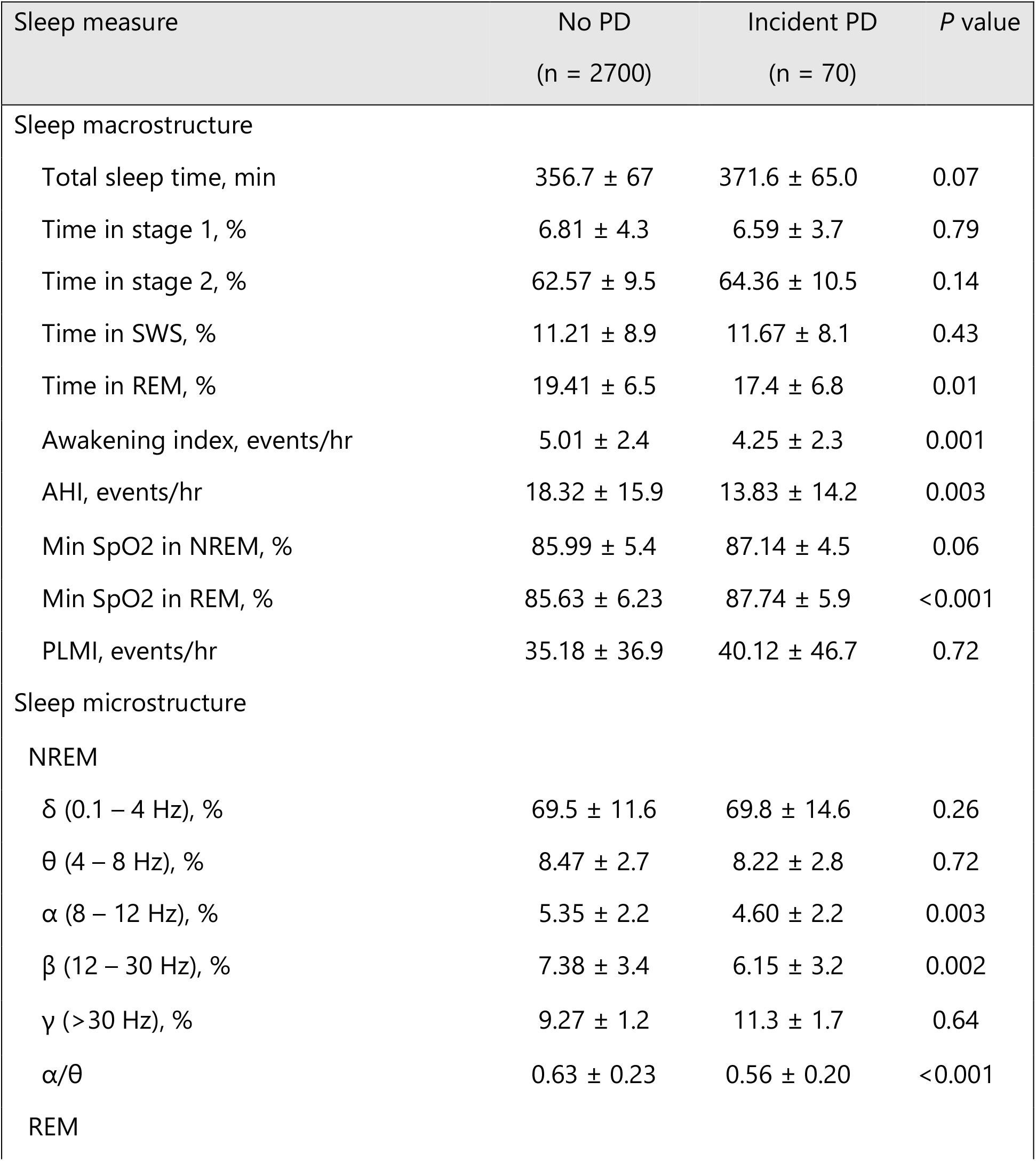
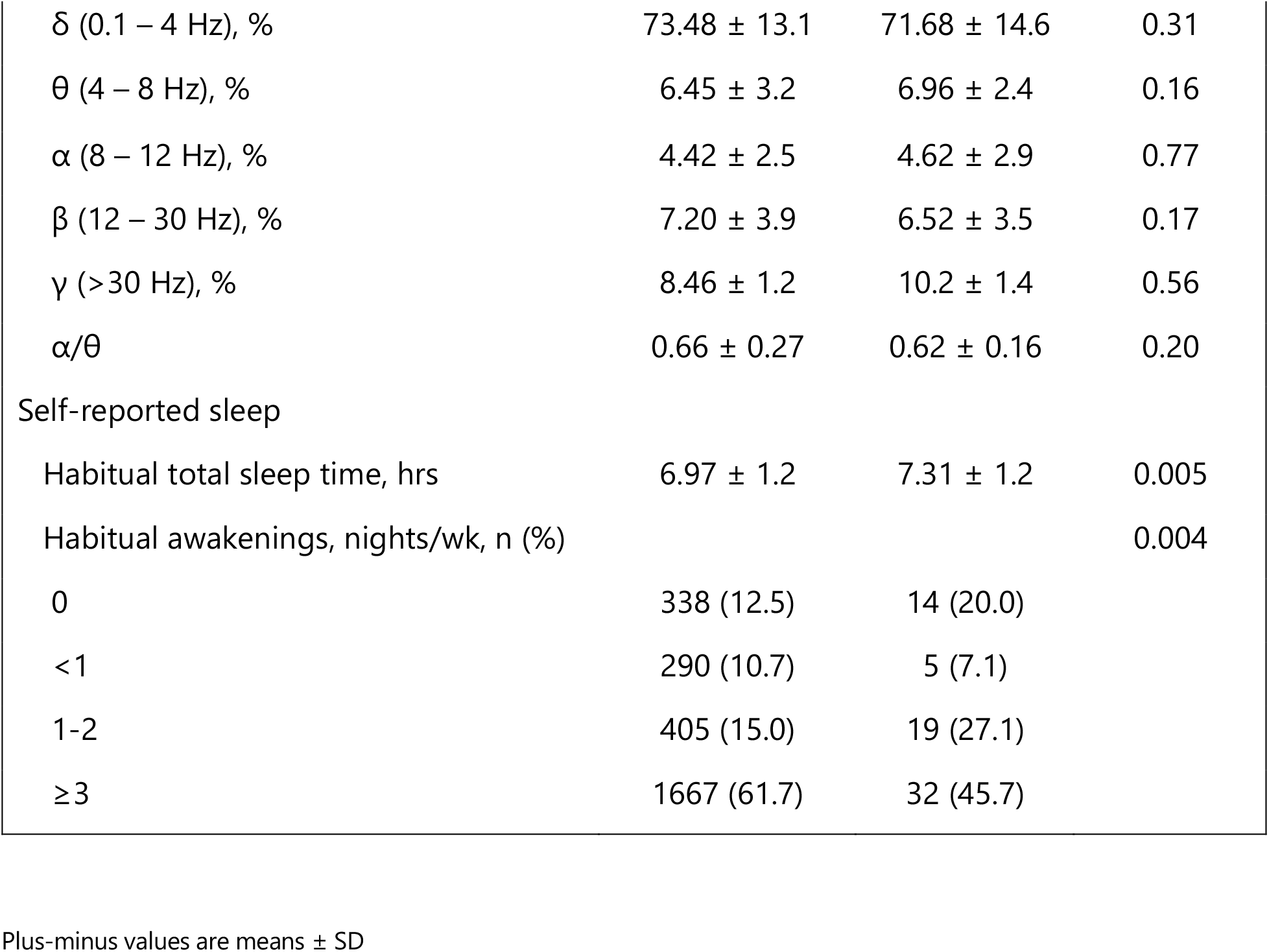
Sleep measures at baseline by incident PD status.

### Sleep macrostructure and risk of developing PD

Lower REM sleep percentage was associated with a higher risk of developing PD across both models. In the fully adjusted model, compared with those in the highest quartile of REM sleep percentage, those in the lowest quartile were 3 times more likely to develop PD (odds ratio [OR], 2.6; 95% CI, 1.3-5.3, *P* = 0.006), and there was a linear trend across quartiles (*P* for trend = 0.01) (Table 3). Stages 1, 2 and SWS were not significantly associated with PD risk. Among the other routine PSG measures, longer TST (OR, 2.2; 95% CI, 1.0-4.5, *P* = 0.046) (P for trend =0.03), and higher min SpO2 during REM sleep (OR, 3.2; 95% CI, 1.5-7.2, *P* = 0.004) (*P* for trend <0.001), were both associated with increased risk of developing PD (Table 3). Conversely, a higher awakening index (OR, 0.35; 95% CI, 0.2-0.8, *P* = 0.006) (P for trend=0.004) and higher AHI (OR, 0.41; 95% CI, 0.2-0.9, *P* = 0.018) (*P* for trend = 0.01), were both associated with decreased risk of developing PD (Table 3).

**Table 3:** Odds ratios and 95% CI for incident PD by sleep macro-structure measure and quartile.

|  |  |  |  |  |  |
| --- | --- | --- | --- | --- | --- |
| **Stage 1, %** |  |  |  |  |  |
|  | Q1 (<4.00) | Q2 (4.00-5.92) | Q3 (5.93-8.57) | Q4 (8.58+) | *P* for trend |
| PD cases [n (%)] | 21 (3.1) | 15 (2.2) | 16 (2.3) | 18 (2.6) |  |
| N | 688 | 697 | 692 | 693 |  |
| Age + clinic adjusted | 1.2 (0.6,2.1) | 0.80 (0.4, 1.6) | 0.87 (0.4, 1.7) | 1 [reference] | 0.77 |
| Multivariate adjusted <sup>a</sup> | 1.1 (0.6,2.0) | 0.8 (0.4, 1.7) | 0.9 (0.4, 1.7)** | 1 [reference] | 0.98 |
| **Stage 2, %** |  |  |  |  |  |
|  | Q1 (<56.40) | Q2 (56.40-62.89) | Q3 (62.89-69.39) | Q4 69.40+) | *P* for trend |
| PD cases [n (%)] | 19 (2.7) | 10 (1.5) | 18 (2.6) | 23 (3.3) |  |
| N | 691 | 689 | 696 | 694 |  |
| Age + clinic adjusted | 0.83 (0.4,1.5) | 0.44 (0.2, 0.9)* | 0.79 (0.4, 1.5) | 1 [reference] | 0.30 |
| Multivariate adjusted <sup>a</sup> | 0.9 (0.5,1.7) | 0.5 (0.2, 1.01) | 0.9 (0.4, 1.6) | 1 [reference] | 0.45 |
| **SWS, %** |  |  |  |  |  |
|  | Q1 (<3.77) | Q2 (3.77-9.92) | Q3 (9.93-16.59) | Q4 (16.60+) | *P* for trend |
| PD cases [n (%)] | 15 (2.2) | 18 (2.6) | 18 (2.6) | 19 (2.7) |  |
| N | 692 | 693 | 685 | 700 |  |
| Age + clinic adjusted | 0.80 (0.4, 1.6) | 0.94 (0.5, 1.8) | 0.98 (0.5, 1.9) | 1 [reference] | 0.53 |
| Multivariate adjusted <sup>a</sup> | 0.7 (0.4,1.6) | 0.9 (0.5, 1.8) | 1.0 (0.5, 2.0) | 1 [reference] | 0.45 |
| **REM, %** |  |  |  |  |  |
|  | Q1 (<14.90) | Q2 (14.90-19.59) | Q3 (19.60-23.69) | Q4 (23.70+) | *P* for trend |
| PD cases [n (%)] | 30 (4.3) | 12 (1.8) | 16 (1.3) | 12 (1.7) |  |
| N | 691 | 680 | 698 | 701 |  |
| Age + clinic adjusted | 2.7 (1.3, 5.7)** | 1.1 (0.5, 2.4) | 1.4 (0.7, 3.0) | 1 [reference] | 0.004** |
| Multivariate adjusted <sup>a</sup> | 2.6 (1.3,5.3)** | 1.0 (0.5, 2.4) | 1.5 (0.7, 3.2) | 1 [reference] | 0.01* |
| **TST, mins** |  |  |  |  |  |
|  | Q1 (<318) | Q2 (318-360) | Q3 (361-400) | Q4 (401+) | *P* for trend |
| PD cases [n (%)] | 11 (1.6) | 16 (2.3) | 20 (2.9) | 23 (3.3) |  |
| N | 690 | 692 | 693 | 695 |  |
| Age + clinic adjusted | 1 [reference] | 1.4 (0.6, 3.1) | 1.8 (0.8, 3.8) | 2.0 (0.97, 4.2) | 0.046* |
| Multivariate adjusted <sup>a</sup> | 1 [reference] | 1.4 (0.6, 3.1) | 1.9 (0.9, 4.1) | 2.1 (1.0, 4.5)* | 0.03* |
| **Awakening index** |  |  |  |  |  |
|  | Q1 (<3.37) | Q2 (3.37-4.53) | Q3 (4.54-5.98) | Q4 (5.99+) | *P* for trend |
| PD cases [n (%)] | 30 (4.4) | 15 (2.2) | 15 (2.1) | 10 (1.4) |  |
| N | 684 | 693 | 700 | 693 |  |
| Age + clinic adjusted | 1 [reference] | 0.49 (0.3, 0.9)* | 0.48 (0.3, 0.9)* | 0.33 (0.2, 0.7)* | 0.002** |
| Multivariate adjusted <sup>a</sup> | 1 [reference] | 0.54 (0.3, 1.0) | 0.54 (0.3, 1.0) | 0.35 (0.2, 0.8)** | 0.004** |
| **AHI** |  |  |  |  |  |
|  | Q1 (<6.30) | Q2 (6.30-13.46) | Q3 (13.47-25.37) | Q4 (25.38+) | *P* for trend |
| PD cases [n (%)] | 28 (4.0) | 16 (2.3) | 14 (2.0) | 12 (1.7) |  |
| N | 692 | 693 | 695 | 690 |  |
| Age + clinic adjusted | 1 [reference] | 0.58 (0.3, 1.1) | 0.51 (0.3, 0.9)* | 0.47 (0.2, 0.9)* | 0.02* |
| Multivariate adjusted <sup>a</sup> | 1 [reference] | 0.60 (0.3, 1.1) | 0.50 (0.3, 1.0)* | 0.41 (0.2, 0.9)* | 0.01* |
| **Min SpO2, % (NREM)** |  |  |  |  |  |
|  | Q1 (<84) | Q2 (84-86) | Q3 (87-89) | Q4 (90+) | *P* for trend |
| PD cases [n (%)] | 11 (1.7) | 15 (2.5) | 22 (2.7) | 22 (3.2) |  |
| N | 643 | 606 | 828 | 693 |  |
| Age + clinic adjusted | 1 [reference] | 1.4 (0.6, 3.0) | 1.5 (0.7, 3.1) | 1.7 (0.8, 3.7) | 0.16 |
| Multivariate adjusted <sup>a</sup> | 1 [reference] | 1.5 (0.7, 3.3) | 1.7 (0.8, 3.6) | 2.1 (0.9, 4.5) | 0.067 |
| **Min SpO2, %(REM)** |  |  |  |  |  |
|  | Q1 (<83) | Q2 (83-86) | Q3 (87-89) | Q4 (90+) | *P* for trend |
| PD cases [n (%)] | 10 (1.5) | 9 (1.5) | 20 (2.9) | 31 (3.8) |  |
| N | 667 | 613 | 681 | 809 |  |
| Age + clinic adjusted | 1 [reference] | 0.9 (0.4, 2.3) | 1.9 (0.9, 4.0) | 2.5 (1.2, 5.2)* | 0.003** |
| Multivariate adjusted <sup>a</sup> | 1 [reference] | 1.1 (0.4, 2.7) | 2.3 (1.0, 5.0)* | 3.2 (1.5, 7.2)** | < 0.001*** |
| **PLMI** |  |  |  |  |  |
|  | Q1 (<3.19) | Q2 (3.19-23.71) | Q3 (23.72-55.56) | Q4 (55.57+) | *P* for trend |
| PD cases [n (%)] | 17 (2.5) | 17 (2.5) | 16 (2.3) | 20 (2.9) |  |
| N | 693 | 692 | 693 | 692 |  |
| Age + clinic adjusted | 1 [reference] | 1.0 (0.51, 2.0) | 0.9 (0.5, 1.9) | 1.1 (0.6, 2.3) | 0.64 |
| Multivariate adjusted <sup>a</sup> | 1 [reference] | 1.0 (0.5, 2.1) | 0.87 (0.4, 1.8) | 1.1(0.6, 2.1) | 0.92 |

Abbreviations: PD, Parkinson's disease; Q, quartile; TST, total sleep time; SWS, slow wave sleep; REM, rapid eye movement sleep; NREM, non-rapid eye movement sleep AHI, apnoea-hypopnea index; min SpO<sub>2</sub>, minimum oxygen saturations; PLMI, periodic limb movement index.
<sup>a</sup> Adjusted for age, clinic site, ethnicity, education, smoking status, diabetes, hypertension, depressive symptoms, cognitive function, daytime sleepiness, physical activity levels, body mass index, caffeine intake, and psychotropic medication use.
\* $P \leq 0.05$
\*\* $P < 0.01$
\*\*\* $P < 0.001$

### Sleep microstructure and risk of developing PD

The strongest association with PD risk was a lower α/θ in NREM sleep. Compared with those in the highest quartile of NREM α/θ, those in the lowest quartile were 4 times more likely to develop PD (OR, 3.7; 95% CI, 1.8-7.7, P <0.001) and there was a linear trend across quartiles (*P* for trend *P* < 0.001) (Table 4). Lower relative α in NREM (OR, 2.3; 95% CI, 1.1-4.8, P=0.03) (*P* for trend *P* = 0.01), and lower relative β in NREM (OR, 2.3; 95% CI, 1.1-4.8, *P* = 0.03) (*P* for trend = 0.01), were also associated with higher risk of incident PD. There were no associations between any qEEG measures during REM sleep, and risk of incident PD (Table 4).

**Table 4:** Odds ratios and 95% CI for incident PD by sleep micro-structure measure and quartile.

| **δ, % (NREM)** |  |  |  |  |  |
| --- | --- | --- | --- | --- | --- |
|  | Q1 (<65.0) | Q2 (65.0-71.6) | Q3 (71.7-76.8) | Q4 (76.9+) | *P* for trend |
| PD cases [n (%)] | 15 (2.2) | 17 (2.5) | 17 (2.5) | 21 (3.0) |  |
| N | 693 | 692 | 692 | 693 |  |
| Age + clinic adjusted | 0.67 (0.3, 1.4) | 0.79 (0.4, 1.5) | 0.80 (0.4, 1.5) | 1 [reference] | 0.28 |
| Multivariate adjusted<sup>a</sup> | 0.72 (0.4, 1.5) | 0.83 (0.4, 1.6) | 0.81 (0.4, 1.6) | 1 [reference] | 0.40 |
| **θ, % (NREM)** |  |  |  |  |  |
|  | Q1 (<6.8) | Q2 (6.8-8.3) | Q3 (8.4-9.9) | Q4 (10.0+) | *P* for trend |
| PD cases [n (%)] | 19 (2.7) | 17 (2.5) | 17 (2.5) | 17 (2.5) |  |
| N | 692 | 693 | 693 | 692 |  |
| Age + clinic adjusted | 1.2 (0.6, 2.4) | 1.1 (0.5, 2.1) | 1.1 (0.5, 2.1) | 1 [reference] | 0.58 |
| Multivariate adjusted<sup>a</sup> | 1.1 (0.6, 2.3) | 1.0 (0.5, 2.0) | 1.0 (0.5, 2.1) | 1 [reference] | 0.74 |
| **α, % (NREM)** |  |  |  |  |  |
|  | Q1 (<3.8) | Q2 (3.8-5.0) | Q3 (5.1-6.4) | Q4 (6.5+) | *P* for trend |
| PD cases [n (%)] | 25 (3.6) | 19 (2.7) | 15 (2.2) | 11 (1.6) |  |
| N | 693 | 692 | 692 | 693 |  |
| Age + clinic adjusted | 2.5 (1.2, 5.1)* | 1.9 (0.9, 4.1) | 1.4 (0.6, 3.1) | 1 [reference] | 0.007** |
| Multivariate adjusted<sup>a</sup> | 2.3 (1.1, 4.8)* | 1.8 (0.8, 3.8) | 1.3 (0.6, 2.8) | 1 [reference] | 0.01* |
| **β, % (NREM)** |  |  |  |  |  |
|  | Q1 (<5.0) | Q2 (5.0-6.6) | Q3 (6.7-9.0) | Q4 (9.1+) | *P* for trend |
| PD cases [n (%)] | 27 (3.9) | 18 (2.6) | 12 (1.7) | 13 (1.9) |  |
| N | 693 | 692 | 692 | 693 |  |
| Age + clinic adjusted | 2.4 (1.1, 5.0)* | 1.6 (0.8, 3.4) | 1.1 (0.5, 2.4) | 1 [reference] | 0.009** |
| Multivariate adjusted<sup>a</sup> | 2.3 (1.1, 4.8)* | 1.5 (0.7, 3.2) | 1.0 (0.4, 2.3) | 1 [reference] | 0.01* |
| **γ, % (NREM)** |  |  |  |  |  |
|  | Q1 (<3.9) | Q2 (3.9-5.7) | Q3 (5.8-9.0) | Q4 (9.1+) | *P* for trend |
| PD cases [n (%)] | 20 (2.9) | 20 (2.9) | 13 (1.9) | 17 (2.5) |  |
| N | 692 | 693 | 692 | 693 |  |
| Age + clinic adjusted | 1.2 (0.6, 2.4) | 1.2 (0.6, 2.4) | 0.79 (0.4, 1.7) | 1 [reference] | 0.38 |
| Multivariate adjusted<sup>a</sup> | 1.2 (0.6, 2.4) | 1.2 (0.6, 2.3) | 0.77 (0.4, 1.6) | 1 [reference] | 0.43 |
| **α/θ (NREM)** |  |  |  |  |  |
|  | Q1 (<0.50) | Q2 (0.50-0.59) | Q3 (0.60-0.71) | Q4 (0.72+) | *P* for trend |
| PD cases [n (%)] | 35 (5.1) | 15 (2.2) | 10 (1.4) | 10 (1.4) |  |
| N | 692 | 693 | 693 | 692 |  |
| Age + clinic adjusted | 3.7 (1.8, 7.7)*** | 1.5 (0.7, 3.4) | 0.96 (0.4, 2.3) | 1 [reference] | <0.001*** |
| Multivariate adjusted<sup>a</sup> | 3.7 (1.8, 7.7)*** | 1.5 (0.7, 3.4) | 0.96 (0.4, 2.3) | 1 [reference] | <0.001*** |
| **δ, % (REM)** |  |  |  |  |  |
|  | Q1 (<67.0) | Q2 (67.0-75.6) | Q3 (75.7-82.5) | Q4 (82.6+) | *P* for trend |
| PD cases [n (%)] | 12 (3.0) | 18 (2.6) | 15 (2.2) | 16 (2.3) |  |
| N | 692 | 693 | 692 | 693 |  |
| Age + clinic adjusted | 1.1 (0.6, 2.2) | 0.99 (0.5, 2.0) | 0.78 (0.4, 1.9) | 1 [reference] | 0.66 |
| Multivariate adjusted<sup>a</sup> | 1.2 (0.6, 2.5) | 1.1 (0.5, 2.2) | 0.96 (0.5, 2.0) | 1 [reference] | 0.49 |
| **θ, % (REM)** |  |  |  |  |  |
|  | Q1 (<4.3) | Q2 (4.3-5.9) | Q3 (6.0-7.8) | Q4 (7.9+) | *P* for trend |
| PD cases [n (%)] | 12 (1.7) | 17 (2.5) | 18 (2.6) | 23 (3.3) |  |
| N | 692 | 693 | 692 | 693 |  |
| Age + clinic adjusted | 0.58 (0.3, 1.2) | 0.85 (0.4, 1.7) | 0.88 (0.5, 1.7) | 1 [reference] | 0.17 |
| Multivariate adjusted<sup>a</sup> | 0.55 (0.3, 1.2) | 0.80 (0.4, 1.6) | 0.90 (0.5, 1.7) | 1 [reference] | 0.12 |
| **α, % (REM)** |  |  |  |  |  |
|  | Q1 (<2.7) | Q2 (2.7-3.8) | Q3 (3.9-5.5) | Q4 (5.6+) | *P* for trend |
| PD cases [n (%)] | 20 (2.9) | 14 (2.0) | 13 (1.9) | 23 (3.3) |  |
| N | 692 | 693 | 692 | 693 |  |
| Age + clinic adjusted | 1.0 (0.5, 1.9) | 0.70 (0.4, 1.4) | 0.61 (0.3, 1.2) | 1 [reference] | 0.90 |
| Multivariate adjusted<sup>a</sup> | 0.93 (0.5, 1.8) | 0.67 (0.3, 1.3) | 0.57 (0.3, 1.1) | 1 [reference] | 0.89 |
| **β, % (REM)** |  |  |  |  |  |
|  | Q1 (<4.4) | Q2 (4.4-6.4) | Q3 (6.5-9.0) | Q4 (9.1+) | *P* for trend |
| PD cases [n (%)] | 20 (2.9) | 18 (2.6) | 19 (2.7) | 13 (1.9) |  |
| N | 693 | 692 | 692 | 693 |  |
| Age + clinic adjusted | 1.7 (0.8, 3.6) | 1.6 (0.8, 3.3) | 1.5 (0.8, 3.2) | 1 [reference] | 0.16 |
| Multivariate adjusted<sup>a</sup> | 1.6 (0.8, 3.4) | 1.5 (0.7, 3.2) | 1.6 (0.8, 3.3) | 1 [reference] | 0.26 |
| **γ, % (REM)** |  |  |  |  |  |
|  | Q1 (<3.1) | Q2 (3.1-4.8) | Q3 (4.9-7.8) | Q4 7.9+) | *P* for trend |
| PD cases [n (%)] | 14 (2.0) | 20 (2.9) | 17 (2.5) | 19 (2.7) |  |
| N | 693 | 692 | 692 | 693 |  |
| Age + clinic adjusted | 0.79 (0.4, 1.6) | 1.1 (0.6, 32.1) | 0.93 (0.5, 21.8) | 1 [reference] | 0.67 |
| Multivariate adjusted<sup>a</sup> | 0.75 (0.4, 1.5) | 1.1 (0.6, 2.1) | 0.94 (0.5, 1.9) | 1 [reference] | 0.56 |

| **$\alpha/\theta$ (REM)** |  |  |  |  |  |
| --- | --- | --- | --- | --- | --- |
|  | Q1 (<0.53) | Q2 (0.53-0.61) | Q3 (0.62-0.73) | Q4 (0.74+) | *P* for trend |
| PD cases [n (%)] | 21 (3.0) | 18 (2.6) | 15 (2.2) | 16 (2.3) |  |
| N | 693 | 692 | 693 | 692 |  |
| Age + clinic adjusted | 1.4 (0.7, 2.8) | 1.3 (0.6, 2.5) | 0.95 (0.5, 1.9) | 1 [reference] | 0.21 |
| Multivariate adjusted<sup>a</sup> | 1.4 (0.7, 2.7) | 1.2 (0.6, 2.5) | 0.92 (0.4, 1.9) | 1 [reference] | 0.28 |

Abbreviations: PD, Parkinson's disease; Q, quartile; REM, rapid eye movement sleep; NREM, non-rapid eye movement sleep $\alpha/\theta$ , $\alpha/\theta$ ratio.
<sup>a</sup> Adjusted for age, clinic site, ethnicity, education, smoking status, diabetes, hypertension, depressive symptoms, cognitive function, daytime sleepiness, physical activity levels, body mass index, caffeine intake, and psychotropic medication use.
\**P* < 0.05
\*\**P* < 0.01
\*\*\**P* < 0.001

### Sensitivity analyses

In both the 2-year time lag analysis and the analysis that adjusted for the presence of non-apnoea sleep disorders, the associations between the significant predictors identified in the primary analysis were similar and remained significant (Table 5).

**Table 5:** Sensitivity analyses on the association between objective sleep characteristics and incident PD.

| **REM, %** |  |  |  |  |  |
| --- | --- | --- | --- | --- | --- |
|  | Q1 (<14.90) | Q2 (14.90-19.59) | Q3 (19.60-23.69) | Q4 (23.70+) | *P* for trend |
| 2-year lag | 2.7 (1.3, 5.6)** | 1.1 (0.5, 2.5) | 1.5 (0.7, 3.4) | 1 [reference] | 0.01* |
| NASD<sup>a</sup> | 2.6 (1.3,5.2)** | 1.0 (0.5, 2.4) | 1.5 (0.7, 3.2) | 1 [reference] | 0.01* |
| Mutually adjusted<sup>b</sup> | 4.0 (1.9, 8.5)*** | 1.2 (0.5, 2.9) | 1.9 (0.8, 4.0) | 1 [reference] | < 0.001*** |
| **TST, mins** |  |  |  |  |  |
|  | Q1 (<318) | Q2 (318-360) | Q3 (361-400) | Q4 (401+) | *P* for trend |
| 2-year lag | 1 [reference] | 1.4 (0.6, 3.2) | 1.7 (0.8, 3.7) | 2.1 (1.0, 4.6) | 0.049* |
| NASD<sup>a</sup> | 1 [reference] | 1.4 (0.6, 3.1) | 1.9 (0.9, 4.1) | 2.1 (1.0, 4.5)* | 0.03* |
| Mutually adjusted<sup>b</sup> | 1 [reference] | 1.7 (0.8, 3.8) | 2.5 (1.2, 5.6)* | 2.5 (1.1, 5.5)* | 0.02* |
| **Awakening index** |  |  |  |  |  |
|  | Q1 (<3.37) | Q2 (3.37-4.53) | Q3 (4.54-5.98) | Q4 (5.99+) | *P* for trend |
| 2-year lag | 1 [reference] | 0.46 (0.2, 0.9)* | 0.53 (0.3, 1.0) | 0.35 (0.2, 0.8)** | 0.006** |
| NASD<sup>a</sup> | 1 [reference] | 0.55 (0.3, 1.0) | 0.53 (0.3, 1.0) | 0.35 (0.2, 0.7)** | 0.004** |
| Mutually adjusted<sup>b</sup> | 1 [reference] | 0.56 (0.3, 1.1) | 0.55 (0.3, 1.1) | 0.44 (0.2, 1.0) | 0.03* |
| **AHI** |  |  |  |  |  |
|  | Q1 (<6.30) | Q2 (6.30-13.46) | Q3 (13.47-25.37) | Q4 (25.38+) | *P* for trend |
| 2-year lag | 1 [reference] | 0.59 (0.3, 1.2) | 0.56 (0.3, 1.1) | 0.42 (0.2, 0.9)* | 0.03* |
| NASD<sup>a</sup> | 1 [reference] | 0.60 (0.3, 1.1) | 0.50 (0.3, 1.0)* | 0.41 (0.2, 0.9)* | 0.01* |
| Mutually adjusted<sup>b</sup> | 1 [reference] | 0.69 (0.3, 1.4) | 0.68 (0.3, 1.4) | 0.66 (0.3, 1.6) | 0.28 |
| **Min SpO<sub>2</sub>, % (REM)** |  |  |  |  |  |
|  | Q1 (<83) | Q2 (83-86) | Q3 (87-89) | Q4 (90+) | Q1 (<83) |
| 2-year lag | 1 [reference] | 1.3 (0.5, 3.4) | 2.4 (1.0, 5.7) | 3.4 (1.8, 8.0)** | 0.002** |
| NASD<sup>a</sup> | 1 [reference] | 1.1 (0.4, 2.7) | 2.2 (1.0, 5.0)* | 3.2 (1.5, 7.2)** | < 0.001*** |
| Mutually adjusted<sup>b</sup> | 1 [reference] | 0.9 (0.4, 2.4) | 2.0 (0.8, 4.8) | 2.4 (1.0, 5.8) | 0.02* |
| **α, % (NREM)** |  |  |  |  |  |
|  | Q1 (<3.8) | Q2 (3.8-5.0) | Q3 (5.1-6.4) | Q4 (6.5+) | *P* for trend |
| 2-year lag | 2.4 (1.1, 5.2)* | 2.0 (0.9, 4.4)** | 1.3 (0.5, 2.9)** | 1 [reference] | 0.01* |
| NASD<sup>a</sup> | 2.3 (1.1, 4.8)* | 1.7 (0.8, 3.7) | 1.3 (0.6, 2.8) | 1 [reference] | 0.01* |
| Mutually adjusted<sup>b</sup> | 0.89 (0.4, 2.9) | 0.75 (0.5, 2.8) | 0.99 (0.4, 2.4) | 1 [reference] | 0.81 |
| **β, % (NREM)** |  |  |  |  |  |
|  | Q1 (<5.0) | Q2 (5.0-6.6) | Q3 (6.7-9.0) | Q4 (9.1+) | *P* for trend |
| 2-year lag | 2.7 (1.2, 6.0)* | 1.6 (0.7, 3.6) | 1.2 (0.5, 2.9) | 1 [reference] | 0.01* |
| NASD<sup>a</sup> | 2.3 (1.1, 4.8)* | 1.5 (0.7, 3.2) | 1.0 (0.4, 2.3) | 1 [reference] | 0.02* |
| Mutually adjusted<sup>b</sup> | 1.0 (0.3, 2.7) | 0.89 (0.4, 2.2) | 0.76 (0.3, 1.8) | 1 [reference] | 0.84 |
| **α/θ (NREM)** |  |  |  |  |  |
|  | Q1 (<0.50) | Q2 (0.50-59) | Q3 (0.60-0.71) | Q4 (0.72+) | *P* for trend |
| 2-year lag | 5.0 (2.1, 12)*** | 2.2 (0.9, 5.5) | 1.4 (0.5, 3.7) | 1 [reference] | < 0.001*** |
| NASD<sup>a</sup> | 3.7 (1.8, 7.7)*** | 1.5 (0.7, 3.4) | 0.96 (0.4, 2.3) | 1 [reference] | < 0.001*** |
| Mutually adjusted<sup>b</sup> | 3.0 (1.2, 7.7)* | 1.3 (0.5, 3.4) | 0.94 (0.4, 2.4) | 1 [reference] | 0.006** |

Abbreviations: PD, Parkinson's disease; Q, quartile; REM, rapid eye movement sleep; TST, total sleep time; AHI, apnoea-hypopnea index; min SpO<sub>2</sub>, minimum oxygen saturations; $\alpha/\theta$ , $\alpha/\theta$ ratio; NASD, non-apnoea sleep disorders.
<sup>b</sup> Adjusted for physician-diagnosed non-apnoea sleep disorders + age, clinic site, ethnicity, education, smoking status, diabetes, hypertension, depressive symptoms, cognitive function, daytime sleepiness, physical activity levels, body mass index, caffeine intake, and psychotropic medication use.
<sup>b</sup> Adjusted for the seven other predictors in the table + age, clinic site, ethnicity, education, smoking status, diabetes, hypertension, depressive symptoms, cognitive function, daytime sleepiness, physical activity levels, body mass index, caffeine intake, and psychotropic medication use.
\* $P < 0.05$
\*\* $P < 0.01$
\*\*\* $P < 0.001$

In the analysis that mutually adjusted for all the significant predictors identified in the primary analysis, REM sleep percentage, TST, awakening index, min SpO2 in REM, and NREM α/θ, were each found to be independently associated with incident PD (*P’s* < 0.05) (Table 5). AHI, NREM α, and NREM β, were no longer associated with incident PD (Table 5).

In the sensitivity analysis that further adjusted for resting oxygen saturations, a higher AHI remained significantly associated with a decreased risk for developing PD, and a higher min SpO2 during REM sleep remained significantly associated with an increased risk for developing PD (*P’s* < 0.05).

In the analysis that used self-reported sleep measures as exposures, self-reported habitual TST and self-reported awakenings were both associated with PD risk in the direction identified by the objective PSG measures. Compared with individuals in the lowest quartile of self-reported habitual TST (≤ 6 hours), those in the highest quartile (≥ 9 hours) were 4 times more likely to develop PD (OR, 3.6; 95% CI, 1.5-6.7, *P* = 0.006), with a linear trend across quartiles (*P* for trend = 0.02). Compared with individuals who reported “not during the past month” for the PSQI item on nocturnal awakenings, those who reported “three or more times a week”, were 62% less likely to develop PD (OR, 0.38; 95% CI, 0.20-0.73, P=0.006), and there was a linear trend across the categories (*P* for trend = 0.009).

## Discussion

In this large prospective study of community-dwelling older men without PD, five PSG measured sleep characteristics (macro- and microstructural) were independently associated with the risk of developing PD during a 12-year follow-up. Longer TST, lower REM sleep percentage, lower α/θ during NREM sleep, and higher min SpO2 during REM sleep, were independently associated with an increased risk of developing PD. Conversely, a higher awakening index was independently associated with a decreased risk of developing PD. There was a 2 to 4-fold difference in risk between extreme quartiles for each of these measures in the fully adjusted models.

This is the first study to investigate the association between PSG measured sleep characteristics in community-dwelling older adults and the subsequent development of PD. Three previous studies investigated the association of self-reported sleep characteristics and PD risk. In the Nurses’ Health Study^17^ it was found that longer self-reported habitual TST was associated with an increased risk for developing PD, even after excluding the first 8-years of follow-up. However, in the Rotterdam study, shorter self-reported TST was found to be associated with an increased risk of developing PD within the first 2-years of follow-up (although this association was reversed after excluding individuals with clinically significant depression and anxiety at baseline).^18^ In the US NIH-AARP Diet and Health Study, no association was found between self-reported TST and PD risk.^19^ Importantly, the present study can confirm using objective methods, that TST is associated with PD risk, and that it is longer rather than shorter TST, that is associated with increased risk. The present finding of higher awakenings during sleep being associated with a decreased risk of developing PD is a novel finding, although is in line with the Nurses’ Health Study, which showed that the number of years nurses had participated in rotating night shift work, had an inverse-dose response relationship with developing PD;^17^ suggesting that sleep disruption may protect against the development of PD.

Cross-sectional studies have shown that PD patients exhibit a range of objectively measured sleep abnormalities from the time of diagnosis.^4,5^ In one of the few studies to use PSG to investigate sleep macrostructure in an early stage PD cohort, Breen et al identified that newly diagnosed patients with PD showed distinct sleep stage abnormalities, including reduced percentage time spent in REM sleep and increased percentage time spent in stage 1 sleep.^4^ This study also identified that patients with PD had abnormal sleep-related breathing, including lower AHI and higher min SpO2 during sleep.^4^ The current study therefore extends these findings, by demonstrating that reduced REM sleep percentage, lower AHI and higher min O2 (particularly during REM sleep) may precede the development of clinical PD by several years.

A small number of cross-sectional studies have investigated sleep microstructure in patients with PD using qEEG,^5^ and have found abnormalities in REM and NREM sleep. However, a recent longitudinal study identified that qEEG alterations in NREM sleep specifically, were associated with faster motor progression in patients with PD over a mean observation time of approximately 5-years.^20^ In addition, experimental evidence using mouse models demonstrated that modulation of NREM qEEG activity may reduce α-synuclein aggregation.^21^ As such, these studies may contextualise the finding that qEEG abnormalities in NREM sleep, rather than REM sleep, were associated with increased risk of incident PD in the present study. In the present spectral analysis, reduced α, β, and α/θ, were all found to be associated with increased PD risk (consistent with the characteristic slowing of EEG activity seen in PD during wakefulness).^10^ However, only the α/θ was independently associated with developing PD when the measures were included in the model together. This suggests that the α/θ may be the strongest qEEG measure to discriminate patients with PD from healthy controls, as is observed with Alzheimer’s disease. ^22^

The finding that lower AHI and higher min SpO2 during REM sleep were associated with increased risk of PD may seem counterintuitive, given the well-established links between obstructive sleep apnoea (OSA) and dementia, as well as additional evidence suggesting patients diagnosed with OSA may have an increased risk of PD.^23^ However, the evidence linking OSA to increased PD risk is inconsistent. In fact, a meta-analysis found that OSA is actually less common in PD than in the general population.^24^ Intriguingly, a recent autopsy study involving Japanese American men from the Honolulu-Asia Aging study, identified that Lewy bodies were less common in people with a greater percentage of sleep time with a SpO2 <95%.^25^ The authors reported an inverse dose-response association, which is consistent with the inverse dose-response association found in the present study with min SpO2 in REM sleep and physician-diagnosed PD. Why oxygen saturations during REM, but not NREM sleep would be associated with PD risk, is not easy to explain, however a recent study identified that community-dwelling adults with RBD exhibited a lower AHI during REM but not NREM sleep.^26^ This suggests that this intriguing finding was not spurious.

By the time a patient presents to their physician with the early symptoms of PD, they will have irreversibly lost a significant proportion of their dopaminergic neurones. It is thus widely accepted that identifying patients at the preclinical stage of PD,^27^ where minimal neurodegenerative changes have occurred, would be the optimum time to provide disease modifying interventions. However, this strategy requires objective biomarkers that could be used on a population-level to identify individuals that may be at high risk for developing PD. Unfortunately, there are currently no approved objective biomarkers to identify PD risk in the general population.^27^ In this study, it is demonstrated that a single-night, in-home, unattended PSG recording, using a low-density EEG montage and finger pulse oximeter, provided five objective biomarkers of PD risk. As such, this study suggests that PSG may be a useful approach to screen for preclinical PD.

### Strengths and Limitations

This study has several strengths including the prospective design, long follow-up period, inclusion of a wide range of potential confounders and the novelty of the approach (this is the first study to investigate the association between PSG measured sleep characteristics and the risk of PD in a large population of community-dwelling adults). Though the study does have limitations. This includes relying on a self-reported physician diagnosis to determine incident PD, which may have led to a misclassification of some cases. Second, it is difficult to exclude the possibility of reverse causality, with objective sleep alterations being early signs of undiagnosed PD, rather than risk factors for developing PD. However, the long follow-up period coupled with the findings from the 2-year time lag analysis, suggest that these abnormalities preceded the development of clinical PD. The sleep measures were quantified using a single-night of PSG; thus, it is possible that a first-night effect may have biased the results. However, given that the associations between longer TST, decreased awakenings and incident PD, were similar when using either self-reported estimates during the previous month, or the single-night PSG measure, suggests the sleep study protocol did not bias the results. Moreover, another study which used the same PSG protocol as MrOS found no evidence of a first-night effect.^28^ Finally, the findings from this study might not be generalisable to women and younger adults.

## Conclusions

In conclusion, longer TST, lower REM sleep percentage, lower α/θ during NREM sleep and higher min SpO2 during REM sleep, were independently associated with an increased risk for developing PD over a 12-year follow-up in community-dwelling older men. A higher awakening index was independently associated with a decreased risk for developing PD. These novel findings demonstrate that objective sleep alterations precede the development of PD by several years, and that a single-night, unattended, in-home PSG assessment, may be a useful way by which to identify individuals in the general population at high-risk of developing PD. Future studies are needed to establish whether these abnormalities are markers of preclinical PD or causal risk factors.

## Data Availability

All data produced in the present work are contained in the manuscript

## Acknowledgements

I thank Professor Roger Barker and Dr David Breen for helpful comments on this article. The Osteoporotic Fractures in Men (MrOS) Study is supported by National Institutes of Health funding. The following institutes provide support: the National Institute on Aging (NIA), the National Institute of Arthritis and Musculoskeletal and Skin Diseases (NIAMS), the National Center for Advancing Translational Sciences (NCATS), and NIH Roadmap for Medical Research under the following grant numbers: U01 AG027810, U01 AG042124, U01 AG042139, U01 AG042140, U01 AG042143, U01 AG042145, U01 AG042168, U01 AR066160, R01 AG066671, and UL1 TR000128. The National Heart, Lung, and Blood Institute (NHLBI) provides funding for the MrOS Sleep ancillary study “Outcomes of Sleep Disorders in Older Men” under the following grant numbers: R01 HL071194, R01 HL070848, R01 HL070847, R01 HL070842, R01 HL070841, R01 HL070837, R01 HL070838, and R01 HL070839. The spectral analyses were supported in part by the American College of Chest Physicians Associated Subspecialty Professors Geriatrics Development Award (PI: R Mehra).

## Ethical Approvals

All men provided written informed consent. Each individual site received institutional review board approval before commencement of the study. The present study received approval from the University of Birmingham (Ref No ERN_21-1463).

## Data Availability

Data from MrOS are available at https://mrosonline.ucsf.edu. The analysis dataset for this specific manuscript are also available from the corresponding author upon request.

## References

1. GBD 2016 Neurology Collaborators. Global, regional, and national burden of neurological disorders, 1990-2016: a systematic analysis for the Global Burden of Disease Study 2016. Lancet Neurol. 2019 May;18(5):459–480.

2. Videnovic A, Golombek D. Circadian and sleep disorders in Parkinson’s disease. Exp Neurol. 2013 May;243:45–56.

3. Claassen DO, Josephs KA, Ahlskog JE, et al. REM sleep behavior disorder preceding other aspects of synucleinopathies by up to half a century. Neurology. 2010 Aug 10;75(6):494–9.

4. Breen DP, Vuono R, Nawarathna U, et al. Sleep and circadian rhythm regulation in early Parkinson disease. JAMA Neurol. 2014 May;71(5):589–595.

5. Zahed H, Zuzuarregui JRP, Gilron R, et al. The Neurophysiology of Sleep in Parkinson’s Disease. Mov Disord. 2021 Jul;36(7):1526–1542.

6. Orwoll E, Blank JB, Barrett- et al. Design and baseline characteristics of the osteoporotic fractures in men (MrOS) study--a large observational study of the determinants of fracture in older men. Contemp Clin Trials. 2005 Oct;26(5):569–85.

7. Song Y, Blackwell T, Yaffe K, et al. Relationships between sleep stages and changes in cognitive function in older men: the MrOS Sleep Study. Sleep. 2015 Mar 1;38(3):411–21.

8. Winkelman JW, Blackwell T, Stone K, et al. Associations of Incident Cardiovascular Events With Restless Legs Syndrome and Periodic Leg Movements of Sleep in Older Men, for the Outcomes of Sleep Disorders in Older Men Study (MrOS Sleep Study). Sleep. 2017 Apr 1;40(4)

9. Zhang L, Samet J, Caffo B, et al. Power spectral analysis of EEG activity during sleep in cigarette smokers. Chest. 2008 Feb;133(2):427–32

10. Massa F, Meli R, Grazzini M, et al. Utility of quantitative EEG in early Lewy body disease. Parkinsonism Relat Disord. 2020 Jun;75:70–75.

11. Buysse DJ, Reynolds CF 3rd, Monk TH, et al. The Pittsburgh Sleep Quality Index: a new instrument for psychiatric practice and research. Psychiatry Res. 1989 May;28(2):193–213.

12. Sheikh JI, Yesavage JA. Geriatric Depression Scale (GDS): recent evidence and development of a shorter version. Clin Gerontol. 1986;5(1-2):165-173.

13. Teng EL, Chui HC. The Modified Mini-Mental State (3MS) examination. J Clin Psychiatry. 1987 Aug;48(8):314–8.

14. Johns MW. A new method for measuring daytime sleepiness: the Epworth sleepiness scale. Sleep. 1991 Dec;14(6):540–5.

15. Washburn RA, Smith KW, Jette AM, et al. The Physical Activity Scale for the Elderly (PASE): development and evaluation. J Clin Epidemiol. 1993 Feb;46(2):153–62

16. Barone JJ, Roberts HR. Caffeine consumption. Food Chem Toxicol. 1996 Jan;34(1):119–29.

17. Chen H, Schernhammer E, Schwarzschild MA, et al. A prospective study of night shift work, sleep duration, and risk of Parkinson’s disease. Am J Epidemiol. 2006 Apr 15;163(8):726–30.

18. Lysen TS, Darweesh SKL, Ikram MK, et al. Sleep and risk of parkinsonism and Parkinson’s disease: a population-based study. Brain. 2019 Jul 1;142(7):2013–2022.

19. Gao J, Huang X, Park Y, Hollenbeck A, Blair A, Schatzkin A, Chen H. Daytime napping, nighttime sleeping, and Parkinson disease. Am J Epidemiol. 2011 May 1;173(9):1032–8.

20. Schreiner SJ, Imbach LL, Werth E, et al. Slow-wave sleep and motor progression in Parkinson disease. Ann Neurol. 2019 May;85(5):765–770.

21. Morawska MM, Moreira CG, Ginde VR, et al. Slow-wave sleep affects synucleinopathy and regulates proteostatic processes in mouse models of Parkinson’s disease. Sci Transl Med. 2021 Dec 8;13(623):eabe7099.

22. Özbek Y, Fide E, Yener GG. Resting-state EEG alpha/theta power ratio discriminates early-onset Alzheimer’s disease from healthy controls. Clin Neurophysiol. 2021 Sep;132(9):2019–2031.

23. Chen JC, Tsai TY, Li CY, et al. Obstructive sleep apnea and risk of Parkinson’s disease: a population-based cohort study. J Sleep Res. 2015 Aug;24(4):432–7.

24. Zeng J, Wei M, Li T, Chen W, et al. Risk of obstructive sleep apnea in Parkinson’s disease: a meta-analysis. PLoS One. 2013 Dec 9;8(12):e82091

25. Gelber RP, Redline S, Ross GW, et al. Associations of brain lesions at autopsy with polysomnography features before death. Neurology. 2015 Jan 20;84(3):296–303.

26. Haba-Rubio J, Frauscher B, Marques-Vidal P, et al. Prevalence and determinants of rapid eye movement sleep behavior disorder in the general population. Sleep. 2018 Feb 1;41(2):zsx197.

27. Heinzel S, Berg D, Gasser T, et al. Update of the MDS research criteria for prodromal Parkinson’s disease. Mov Disord. 2019 Oct;34(10):1464–1470.

28. Quan SF, Griswold ME, Iber C, et al. Short-term variability of respiration and sleep during unattended nonlaboratory polysomnography--the Sleep Heart Health Study. [corrected]. Sleep. 2002 Dec;25(8):843-9. Erratum in: Sleep. 2009 Oct 1;32(10):table of contents.

